# Machine Learning and Radiomics for Osteoporosis Risk Prediction Using X-ray Imaging

**DOI:** 10.1101/2022.02.03.22270400

**Authors:** Saba Dadsetan, Gene Kitamura, Dooman Arefan, Yuan Guo, Kadie Clancy, Lu Yang, Shandong Wu

## Abstract

Osteoporosis is a significant health and economic issue, as it predisposes patients to a higher risk of bone fracture. Measuring bone mineral density has been shown to be an accurate way to assess the risk for osteoporosis. The most common way for bone density testing is a dual-energy X-ray absorptiometry (DEXA) scan, which may be recommended for patients with increased risk of osteoporosis. Radiograph imaging is widely available in clinical settings and acquired for many reasons, such as trauma or pain. The goal of this project is to extract radiomics information from pelvic X-rays (both the hip and femoral neck regions) to assess the risk of osteoporosis (triaging patients into “normal” vs. “at-risk”, or “low risk” vs. “high risk” categories). The motivation here is not to replace the DEXA scan but to proactively identify patients at risk for osteoporosis and appropriately refer them to management options. We apply machine learning-based radiomics techniques on a study cohort of 565 patients. Our preliminary results show that a correlation between the radiomics features extracted from pelvic X-rays and the level of osteoporosis risk derived from the DEXA test results.

## 1. Introduction

In the US, almost 44 million people over age 50 are at risk of developing osteoporosis [1]. In 2002, the cost of osteoporosis was estimated at $14 billion [2]. To diagnose this significant health and economic issue at an early stage, there are some available techniques. Various questionaries have been created to identify osteoporosis risk factors, which require patients’ cooperation and input such as family history of osteoporosis, smoking habits, sun exposure, etc. [3]. Following the gold standard method by the World Health Organization (WHO) to assess osteoporosis, measuring bone mineral density (BMD) is recommended via a dual-energy X-rays absorptiometry (DEXA) scan. This method is accurate with moderate cost, but it involves modest radiation exposure. For each patient, the DEXA test results are reported in two numbers: T-score and Z-score [4]. T-score represents a number that compares the condition of the bones with those of an average young person with healthy bone, while Z-score compares the bones with an average same-aged person with healthy bone. Among these two scores, T-score is the most common measurement to consider, particularly for women after menopause and men older than 50 years old [5].

The benefits of the DEXA scan is predicated on the cooperation of patients and availability of the scanning devices. In clinical practice, X-ray and computed tomography (CT) are more common examinations. In prior studies, the CT-based imaging features extracted from chest/abdominal images [6] and spinal images [7] have shown the potentials for osteoporosis fracture risk assessment (i.e., low risk vs. high risk) and for spinal bone mineral density classification (i.e., normal vs. abnormal). Despite the increased availability of CT scans, the associated radiation exposure is much higher than X-ray imaging. X-ray scans are also less costly compared to other imaging modalities and in practice, a significant number of X-ray radiographs are acquired in current clinical settings for various reasons, such as trauma, pain, etc. The purpose of this work is to investigate a machine learning-based computational model for predicting risk of osteopenia and osteoporosis by using radiomics on clinically readily available X-ray images.

## 2. Materials and Methods

### 2.1 Study Cohort and Database

This retrospective study is approved by institutional review board at our institution. We assembled a cohort of 565 patients treated at the University of Pittsburgh Medical Center (UPMC). Pelvic X-ray scans, DEXA scores, and clinical reports are available for all patients in this study cohort. The DEXA scores are available in the form of T-scores for both the hip and neck femoral regions. All the X-ray images of the study cohort were acquired during the year 2010 to 2017. For each patient, the time interval between the X-ray imaging and the DEXA scan is 180 days or less. Demographic information such as sex and age are identified for the full study cohort. Out of the 565 patients, we used the 424 patients (images acquired during January 2010 - April 2014) as the training set, and the remaining 141 patients (images acquired during May 2014 - May 2017) as an independent test set (no overlap with the training set).

### 2.2 Methodology

As shown in **Figure 1**, our method pipeline consists of five major steps: (a) task definition, (b) image annotation, (c) extracting radiomics features, (d) selecting features, and (e) classification.

**Figure 1.**
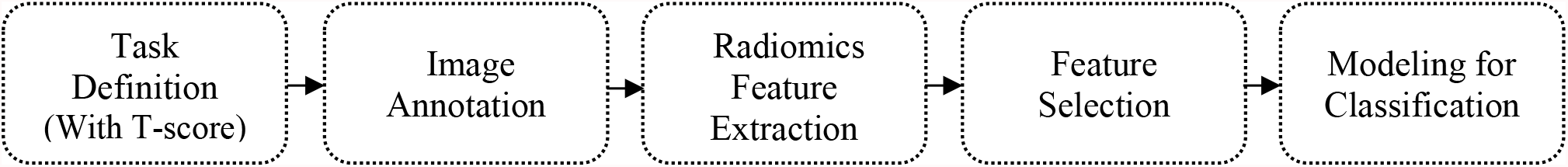
The workflow of osteoporosis risk prediction using radiomics of pelvic X-ray images.

#### 2.2.1 Task Definition

The task is to build an osteoporosis risk prediction model. To that end, we separated the patients into two groups: “normal” or “at-risk” of having osteoporosis. According to the WHO guideline, the lower the T-score, the greater the risk of fracture. Three ranges of the T-score values are characterized: greater than -1, between -1 and -2.5, and lower than -2.5, representing normal, osteopenia, and osteoporosis, respectively. Based on these T-score thresholds, two tasks are defined: I) “normal” vs “at-risk” (osteopenia and osteoporosis) and II) “low risk” (normal and osteopenia) vs “high risk” (osteoporosis). It should be noted that a patient may have different T-scores measured separately at the hip and neck region. To avoid the potential discrepancy between the hip-measured T-score and the neck-measured T-score, based on our expert physicians, we use the lower score of the two T-scores to assign a patient to a classification category. **Table 1** shows the breakdown of patient numbers for the two tasks.

**Table 1.** Number of patients in the training set and independent testing set for the two different tasks.

| Cohort | Task I |  | Task II |  | Total<br>(n=565) |
| --- | --- | --- | --- | --- | --- |
|  | Normal | At-risk | Low risk | High risk |  |
| Training set (2010 to 2014) | 140 | 284 | 341 | 83 | 424 |
| Test set (2014 to 2017) | 45 | 96 | 107 | 34 | 141 |

#### 2.2.2 Image Annotation

In pelvic DEXA scan, BMD may be assessed from several potential regions of interest (ROIs). In our study, we have the T-scores measured separately from the hip and femoral neck regions in the pelvic X-ray. We aim to focus on these two regions as the ROIs to extract radiomics imaging features. To do so, the ROIs of the hip and femoral neck for each patient are manually annotated with bounding boxes (i.e., a rectangle region) by a certified radiologist using the “LabelMe” software [8], a semi-automated software to create a bounding box of a ROI in a given image. Furthermore, for a small subset of 100 X-ray images, another radiologist separately annotated the ROIs, where the Cohen’s kappa statistics [9] is used to measure the inter-reader annotation variability between the two radiologists. **Figure 2** shows an example of the ROI annotations in the hip region (Figure 2 **(b)**) and the femoral neck region (Figure 2 **(c)**).

**Figure 2.**
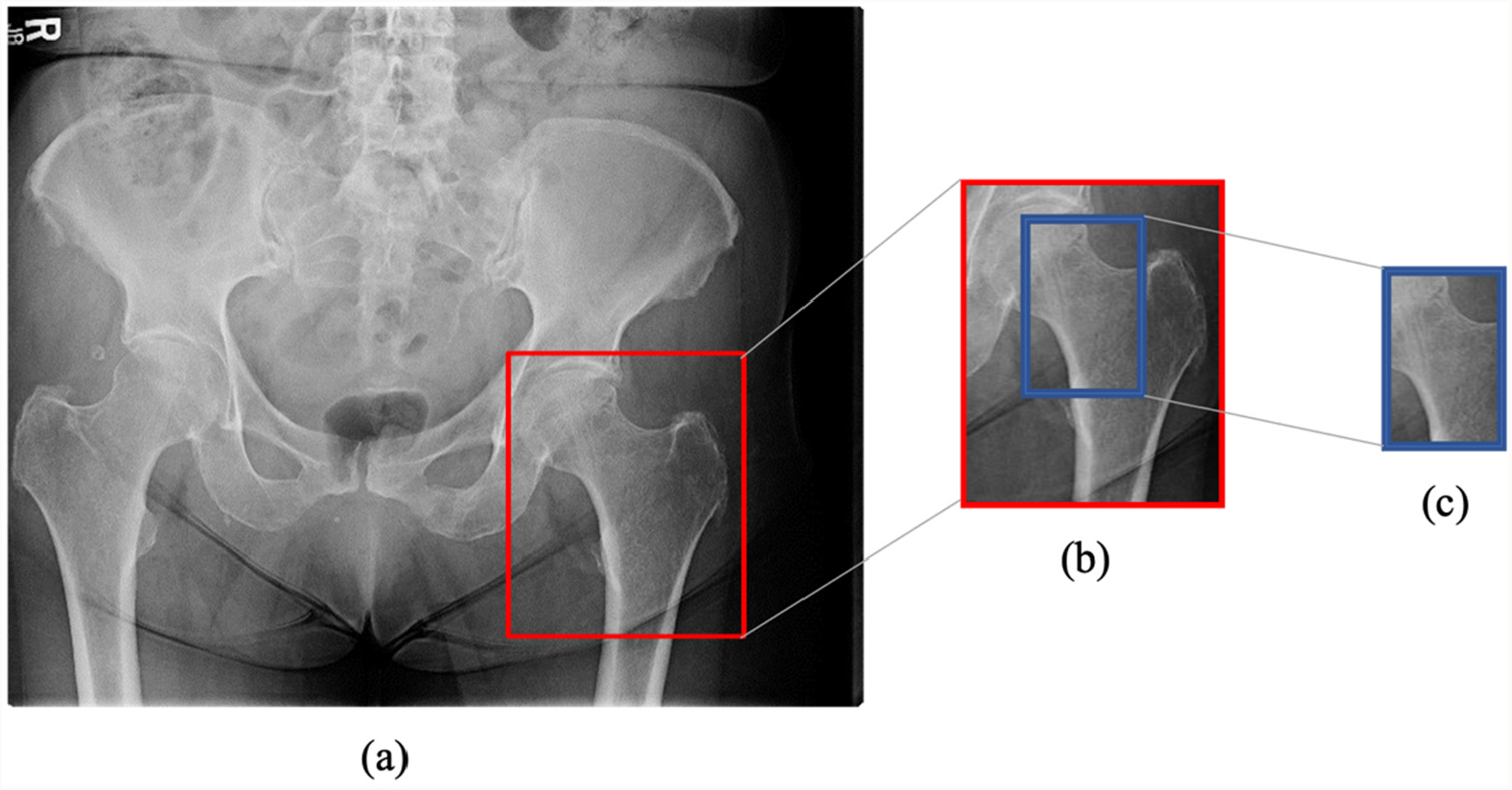
An example of the annotations of the regions of interest for radiomic feature extraction to estimate bone mineral density. (a) X-ray image of the whole pelvis, (b) Annotated region of the left side hip, (c) Annotated region of the left side femoral neck.

Note that before image annotation, all X-ray images are first made consistent in grayscale for anatomical representation (i.e., bones are represented/shown as “bright”). For those images where the bones appear originally “dark” instead of “bright”, we use Python and OpenCV library (the bitwise NOT function) to invert the grayscale so that all the images are made consistent in the representation.

#### 2.2.3 Feature extraction, selection, and modeling for classification

We use radiomics features of the ROIs to identify imaging-related information for BMD estimation. We extract 93 radiomics features [10] from each annotated ROI of the hip or neck regions. These features include first-order statistics of the intensity histogram, 2D geometrics properties of the bone as shape features, textural features derived from the gray level co-occurrence (GLCM) and run-length metrics (GLRLM), gray level size zone matrix (GLSZM), neighboring gray tone difference Matrix (NGTDM), and gray level dependence matrix (GLDM). We use PyRadiomics [11], an open-source python package, to extract the radiomic features from the annotated ROIs. We employ 5 different machine learning models (i.e., logistic regression (LR), random forest (RF), XG-boost, support vector machine (SVM), and k-nearest neighbor (KNN)) for classification. We examine 2 feature selection methods (i.e., LASSO [reference] and Chi-Square) to reduce the dimensionality of the radiomics features. The effects of the feature selection methods and machine learning models are compared on the training set, and we identify the best-performing combination to report the results on the independent test set. The area under the ROC curve (AUC) alongside sensitivity and specificity are used as the model performance metrics. Note that we use the StandardScaler method from the Python scikit-learn package (Version 1.0.2) to standardize the features on the training set and test set. In the training step, we use 5-fold cross-validation to adjust hyperparameters and choose the optimal model. For statistical analysis, we use the SAS software (Version 9.2) to calculate confidence intervals (Mann-Whitney test), p-values, sensitivity at a fixed specificity, and specificity at a fixed sensitivity [12]. P-values < 0.05 are considered as statistically significant. We use the naïve binominal method to estimates the variances (upper and lower limits) of sensitivity and specificity.

## 3. Results

A majority of the patients in the study cohort are female (70%), while 30% are male. The age of the patients ranges from 35 to 91 (mean value: 62 years old). There is a significant (p = 0.02) difference in the mean age between the training and testing set, while the difference between the gender distributions in the training and testing set is insignificant (p = 0.06).

The Cohen’s kappa statistics of the two radiologists in annotating the images on the subset data are 0.96 (mean) with a standard deviation of 0.02 for hip regions, and 0.96 (mean) with a standard deviation of 0.11 for femoral neck regions. These measures indicate a high inter-reader annotation agreement for both regions.

Out of the examined machine learning models and feature selection methods, the best-performing combination is the Chi-Square feature selection and LR classifier, and the corresponding results are reported in the following. **Table 2** shows the AUC values for the two tasks. As can be seen, when comparing radiomics and age+sex, radiomics always underperformed age+sex, except for task II when using the hip region for classification. However, when combining the radiomics and the age+sex variables, it led to increased (but not always statistically significant) AUC values than using the radiomics alone or age+sex alone. This effect is observed for both tasks and for both the hip and neck region. For instance, in task I, in the neck region the combination of features i.e., radiomics + age + sex (AUC = 0.68) achieves a higher AUC than radiomics (AUC = 0.58, p = 0.04) and age + sex (AUC = 0.64, p = 0.38). Likewise, in task II, the combination of features leads to an improved performance in the neck region (AUC = 0.60) compared to radiomics (AUC = 0.54, p = 0.11) and age + sex (AUC = 0.58, p = 0.74). Only in task I, the combination of features shows statistically significant (p<0.05) improvement over the radiomics.

**Table 2.** Model performance for the two tasks of assessing the risk of osteoporosis using different sets of features. Numbers represent AUC values, their 95% confidence intervals (CIs), and p-values. The asterisk indicates statistical significance (p < 0.05) of the AUC differences when compared to the Combination model.

|  |  | Combination<br>(Radiomics + Sex + Age) | Radiomics |  | Sex + Age |  |
| --- | --- | --- | --- | --- | --- | --- |
|  |  | AUC (95% CI) | AUC (95% CI) | p-value | AUC (95% CI) | p-value |
| Task I (“normal” vs “at-risk”) | Hip region | 0.68 (0.58, 0.77) | 0.62 (0.52, 0.72) | 0.03* | 0.64 (0.54, 0.73) | 0.40 |
|  | Neck region | 0.68 (0.59, 0.77) | 0.58 (0.48, 0.69) | 0.04* | 0.64 (0.54, 0.73) | 0.38 |
| Task II (“low risk” vs “high risk”) | Hip region | 0.66 (0.55, 0.76) | 0.63 (0.51, 0.75) | 0.56 | 0.58 (0.46, 0.69) | 0.25 |
|  | Neck region | 0.60 (0.49, 0.71) | 0.54 (0.43, 0.65) | 0.11 | 0.58 (0.46, 0.69) | 0.74 |

**Tables 3** and **4** report further comparisons of the models in terms of sensitivity and specificity. To compare the performance of the two regions, in task I both regions using the combination of features achieve the same AUC of 0.68, however, as shown in **Tables 3** and **4**, the hip region shows the same or higher sensitivities and specificities at the fixed thresholds. This indicates that features of the hip region have a higher correlation to the prediction tasks than the features of the neck region. Interestingly, in task II, all three metrics (AUC, sensitivity, specificity) highlight that the hip region performs better in most cases than the neck region. Furthermore, **Table 4** shows sex+age alone achieves higher specificities at the two fixed sensitivities (i.e., 0.99 and 0.90), compared to the corresponding radiomics. There are no consistent observations for the results shown in **Table 3** between radiomics and age+sex features. Overall, we observe both radiomics and age+sex features contribute to the combination model in achieving the highest performance in both tasks. **Figure 3** shows the ROC curves for four experimental scenarios.

**Table 3.** Sensitivity values and their lower and upper limits for the two tasks at the fixed point of specificity (i.e., 0.99 and 0.90).

|  |  | Task I (“normal” vs “at-risk”) |  | Task II (“low risk” vs “high risk”) |  |
| --- | --- | --- | --- | --- | --- |
|  |  | Hip region | Neck region | Hip region | Neck region |
|  |  | Sensitivity<br>(Lower - Upper) | Sensitivity<br>(Lower - Upper) | Sensitivity<br>(Lower - Upper) | Sensitivity<br>(Lower - Upper) |
| Specificity<br>0.99 | Combination | 0.09 (0.04 - 0.17) | 0.02 (0.002 - 0.07) | 0.17 (0.06 - 0.34) | 0 (0 - 0.1) |
|  | Radiomics | 0.05 (0.01 - 0.10) | 0.01 (0.0002 - 0.05) | 0.23 (0.10 - 0.41) | 0.02 (0.0007 - 0.15) |
|  | Sex+age | 0.04 (0.01 - 0.10) | 0.03 (0.006 - 0.08) | 0.05 (0.007 - 0.19) | 0.05 (0.007 - 0.19) |
| Specificity<br>0.90 | Combination | 0.23 (0.15 - 0.33) | 0.15 (0.09 - 0.24) | 0.26 (0.12 - 0.44) | 0.17 (0.06 - 0.34) |
|  | Radiomics | 0.16 (0.09 - 0.25) | 0.13 (0.07 - 0.22) | 0.35 (0.19 - 0.53) | 0.14 (0.04 - 0.31) |
|  | Sex+age | 0.29 (0.20 - 0.39) | 0.29 (0.20 - 0.39) | 0.29 (0.15 - 0.47) | 0.29 (0.15 - 0.47) |

**Table 4.** Specificity values and their lower and upper limits for the two tasks at the fixed point of sensitivity (i.e., 0.99 and 0.90).

|  |  | Task I (“normal” vs “at-risk”) |  | Task II (“low risk” vs “high risk”) |  |
| --- | --- | --- | --- | --- | --- |
|  |  | Hip region | Neck region | Hip region | Neck region |
|  |  | Specificity<br>(Lower - Upper) | Specificity<br>(Lower - Upper) | Specificity<br>(Lower - Upper) | Specificity<br>(Lower - Upper) |
| Sensitivity<br>0.99 | Combination | 0.06 (0.01 - 0.18) | 0.02 (0.0005 - 0.11) | 0.06 (0.02 - 0.13) | 0 (0 - 0.03) |
|  | Radiomics | 0 (0 - 0.07) | 0 (0 - 0.07) | 0.009 (0.002 - 0.05) | 0.05 (0.02 - 0.11) |
|  | Sex+age | 0.02 (0.0005 - 0.11) | 0.02 (0.0005 - 0.11) | 0.09 (0.04 - 0.16) | 0.09 (0.04 - 0.16) |
| Sensitivity<br>0.90 | Combination | 0.35 (0.21 - 0.51) | 0.26 (0.14 - 0.41) | 0.22 (0.12 - 0.28) | 0.20 (0.13 - 0.29) |
|  | Radiomics | 0.28 (0.16 - 0.44) | 0.17 (0.08 - 0.32) | 0.01 (0.0022 - 0.06) | 0.10 (0.05 - 0.17) |
|  | Sex+age | 0.29 (0.16 - 0.44) | 0.29 (0.16 - 0.44) | 0.13 (0.07 - 0.20) | 0.13 (0.07 - 0.20) |

**Figure 3.**
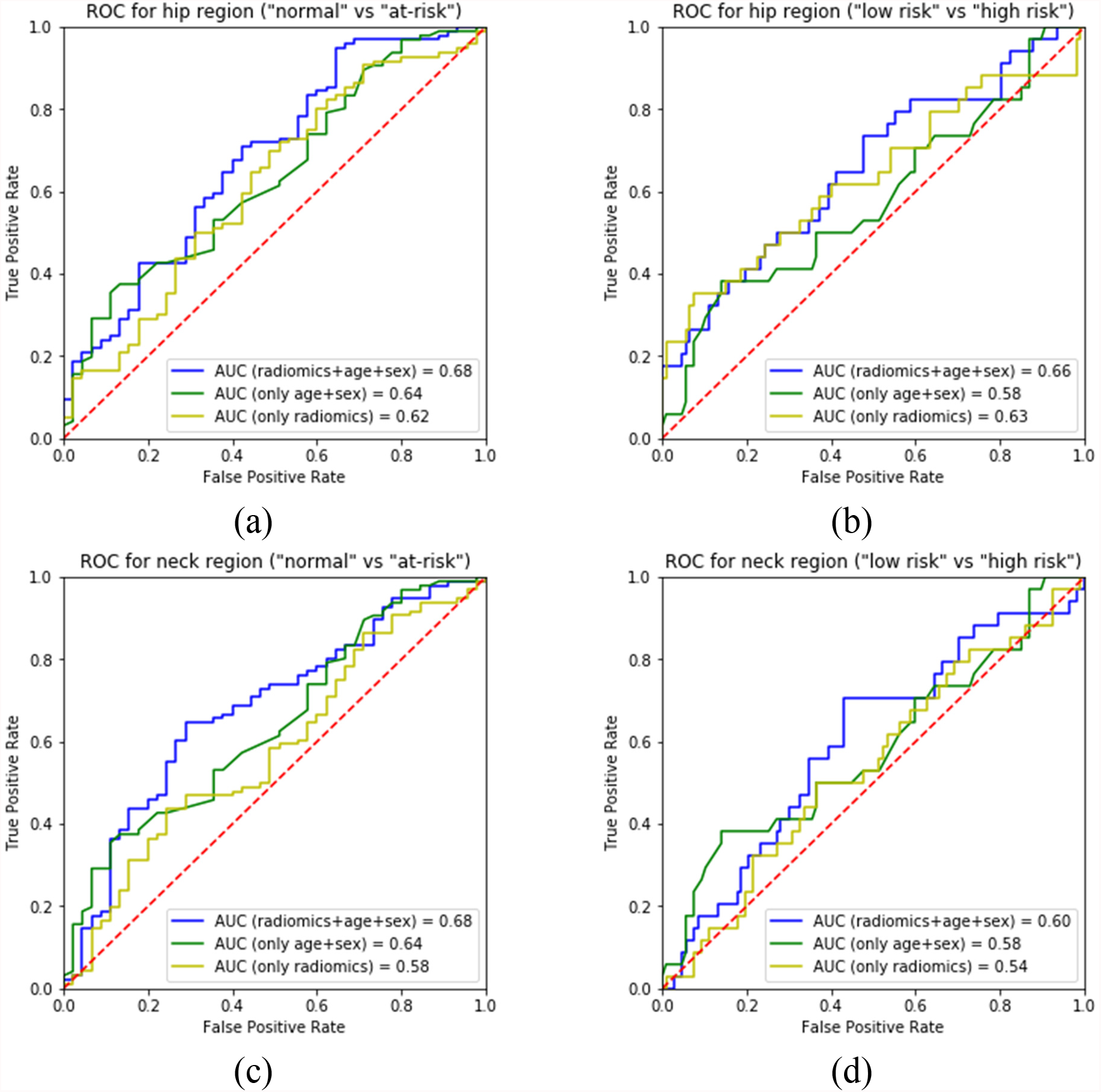
The ROC curves of four experimental scenarios: (a) Task I (“normal” vs “at-risk”) for hip region. (b) Task II (“low risk” vs “high risk”) for hip region. (c) Task I for neck region. (d) Task II for neck region. Each plot shows three ROC curves for using radiomics alone, sex and age, and their combination.

## 4. Discussion

In this study, we took advantage of pre-existing X-ray images to extract radiomics features alongside demographic information (i.e., sex and age) to predict the risk of osteoporosis. We defined two tasks and evaluated the effects of two different ROIs in pelvic images. Our study included a large dataset of 565 patients, and we reported the model performance on the independent test set.

The AUC of our models leaves room for improvement with the highest value of 0.68. Furthermore, there was no statistically significant difference between models based on radiomics vs. radiomics plus demographics. As age and gender are two of the major non-modifiable risk factors for osteoporosis, it follows that a model based only on these two attributes could predict osteoporosis risk to a certain extent [13]. However, it was promising to see that adding in the radiomics feature increased the AUC, although it didn’t reach statistical significance (e.g., 0.64 to 0.68 with a p-value of 0.38 for Task I in the neck region). Our models also appear to stratify normal vs. at-risk better (AUC 0.68) than low-risk vs. high-risk (AUC 0.60-0.66). This result implies that the model would work best at identifying at-risk patients, who then should be referred for further workup.

Examining the sensitivity and specificities at fixed values showed that evaluating the hip region as a whole, generally performed better than being coned down on the femoral neck region. As the hip region encompasses a larger area, it follows that the model was able to perform better when given a larger region to extract radiomics information. Practically, this result is promising, as a unilateral hip X-ray may potentially be used as the input image rather than requiring an intervening bounding box step prior to model application. The fact that demographics performed best with certain metrics and thresholds, reinforced the knowledge that age and sex are major contributors to osteoporosis risk.

The machine learning model-generated estimate of the risk categories may be clinically valuable for practicing physicians and patients at the point of contact. As our output metrics leave room for improvement, it should be pointed out that our goal is not to replace the gold standard DEXA scan, but instead to provide a risk estimate using the analytics on the pre-existing X-rays. We anticipate that these risk estimates could flag a patient at risk and alert the clinician for a potential DEXA referral. In 2018, there were approximately 15,000 pelvic x-rays performed in the health system of our institution, with each case being a potential point of application, leading to potential DEXA tests. Studies have shown that patients undergoing DEXA tests have been estimated to convert to treatment at 44% [14]. By initiating treatment, patients can reduce their 5-year fracture risk from 34% to 10% [15]. This can in turn lead to decreased fracture-related medical costs by 32% and improved patient quality of life [16]. This potential benefit can be realized for the patient without any additional risk, as the risk estimate value is extrapolated from the pre-existing X-ray images.

The limitation of this study is the suboptimal AUC and output metrics value, which may be increased with a higher sample size and more accurate ROI definition. The model performance could be increased with a stricter area of interest on the X-rays to extract radiomics features, possibly going with accurate bone segmentation rather than using a bounding box. We will explore this method in future work.

In summary, our radiomics-based machine learning models show promise in extracting objective osteoporosis risk measures from pre-existing radiographs, creating a potential point of intervention for clinicians in managing the often silent and missed osteoporosis cases. Further investigations are warranted.

## Data Availability

The imaging and patient data in the present study are not available due to privacy considerations. The result data are available upon reasonable request to the authors.

## Acknowledgment

This work was supported in part by the Pilot Research Program of the Pittsburgh Center for AI Innovation in Medical Imaging and the associated Pitt Momentum Funds of a Scaling grant from the University of Pittsburgh (2020) and by an Amazon Machine Learning Research Award.

## Notes

### Competing Interest Statement

Shandong Wu is a scientific consultant and stockholder of COGNISTX, Inc for work unrelated to this study. Shandong Wu receives research grants from the National Cancer Institute, National Institute of Biomedical Imaging and Bioengineering, National Science Foundation, Radiological Society of North America, UPMC Hillman Cancer Center, and Amazon.

### Author Declarations

IRB of the University of Pittsburgh gave ethical approval for this work.

